# Preliminary reference values for Alzheimer’s disease plasma biomarkers in Congolese individuals with and without Alzheimer’s disease

**DOI:** 10.1101/2024.08.06.24311577

**Authors:** Jean Ikanga, Kharine Jean, Priscilla Medina, Saranya Sundaram Patel, Megan Schwinne, Emmanuel Epenge, Guy Gikelekele, Nathan Tshengele, Immaculee Kavugho, Samuel Mampunza, Lelo Mananga, Charlotte E. Teunissen, Anthony Stringer, Julio C. Rojas, Brandon Chan, Argentina Lario Lago, Joel H. Kramer, Adam L. Boxer, Andreas Jeromin, Alden L. Gross, Alvaro Alonso

## Abstract

**Background:** Western countries have provided reference values (RV) for Alzheimer’s disease (AD) plasma biomarkers, but there are not available in Sub-Saharan African populations.

**Objective:** We provide preliminary RV for AD and other plasma biomarkers including amyloid- β (Aβ42/40), phosphorylated tau-181 and 217 (p-tau181, p-tau217), neurofilament light (Nfl), glial fibrillary acidic protein (GFAP), interleukin 1b and 10 (IL-1b and IL-10) and tumor necrosis factor α (TNFα) in Congolese adults with and without dementia.

**Methods:** 85 adults (40 healthy and 45 dementia) over 50 years old were included. Blood samples were provided for plasma AD biomarkers Aβ42/40 and p-tau181, p-tau217; Nfl and GFAP; IL-1b and IL-10 and TNFα analyzed using SIMOA. Linear and logistic regressions were conducted to evaluate differences in biomarkers by age and gender and neurological status, and for the prediction of dementia status by each individual biomarker. RV were those that optimized sensitivity and specificity based on Youden’s index.

**Results:** In this sample of 85 adults, 40 (47%) had dementia, 38 (45.0%) were male, overall mean age was 73.2 (SD 7.6) years with 8.3 (5.4) years of education. There were no significant differences in age, gender, and education based on neurological status. Biomarker concentrations did not significantly differ by age except for p-tau181 and GFAP and did not differ by sex. Preliminary cutoffs of various plasma in pg/ml were 0.061 for Aβ42/40, 4.50 for p-tau 181, 0.008 for p-tau 217, 36.5 for Nfl, 176 for GFAP, 1.16 for TNFa, 0.011 for IL-1b, and 0.38 for IL-10. All AUCs ranged between 0.64-0.74. P-tau 217 [0.74 (0.61, 0.86)] followed by GFAP [0.72 (0.61, 0.83), and Nfl [0.71 (0.60, 0.82)] had the highest AUC compared to other plasma biomarkers.

**Conclusions:** This study provides RV which could be of preliminary utility to facilitate the screening, clinical diagnostic adjudication, classification, and prognosis of AD in Congolese adults.

## INTRODUCTION

Alzheimer’s disease (AD) is a progressive neurodegenerative disorder.[1] Ongoing development in the research of AD pathology has expanded the number of fluid (e.g., cerebral spinal fluid [CSF], plasma) biomarkers recognized in the screening, diagnosis, and monitoring of AD.[2] Current revised 2023 Alzheimer’s Association (AA) criteria differentiates between two broad categories of AD fluid biomarkers related to AD pathogenesis: (1) core AD fluid biomarkers (the CSF ratio of amyloid-β [Aβ42/40], phosphorylated tau-181 and plasma 217 [p- tau181, p-tau217]) and (2) non-specific biomarkers involved in other neurodegenerative pathology, including neurofilament light (Nfl) and glial fibrillary acidic protein (GFAP).[3] Although not included in the AA core-criteria of the final document, neuroinflammatory/immune biomarkers, interleukin 1b and 10 (IL-1b and IL-10) and tumor necrosis factor α (TNFα), play an important role in other neurodegenerative.[2,4–8]

Reference values (RV) of these plasma biomarkers within non-White samples is not yet established, as most studies have been primarily conducted using largely non-Hispanic White (NHW) individuals.[9] There appears to be some evidence of ethnoracial differences in the levels of plasma AD biomarkers and their diagnostic precision based on age and sex,[10–13] emphasizing the need for further research examining these associations. Given the intra- and inter-assay variability, as well as inconsistencies and differences in methods, defining a set of universal cut-offs for plasma AD biomarkers is challenging and may not be possible.[14] It has been recommended that studies define cut-offs in-house to best represent specific populations, with the recognition that the establishment of contemporary clinical cut-offs will need to be assay-dependent.[14,15] Alternatively, calibration equations could be developed to harmonize biomarkers across assays and labs; however, such activities require complicated sample exchanges which can be challenging in cross-national research.

Some studies have established reference intervals, values, and cutoffs based on demographic characteristics such as age and sex. A recent study compared biomarker levels (plasma Aβ42/40, p-tau181, GFAP, and Nfl) by age and gender in participants who were cognitively unimpaired, mildly cognitively impaired, and Aβ-PET–positive participants across the AD pathology continuum.[15] In the AD group, Aβ42/40, p-tau181, and GFAP did not show a significant difference across age; however, older age was associated with higher Nfl concentrations.[15] In a sample of healthy Chinese older adults, Chen and colleagues (2023) identified reference intervals for specific age groups (i.e., 50-59, 60-69, 70-79, 80-89), as well as sex differences. Within their sample, there were no differences between women and men in plasma Aβ42, Aβ40, or Aβ42/40 ratio; however, men had greater plasma p-tau181, p-tau181/t-tau ratio, and p-tau181/Aβ42 ratio than women.[16] In general, age and sex-specific cut-offs for plasma biomarker diagnostic and prognostic use may be important, particularly given the analytical variability and difference in plasma biomarkers’ concentrations across ethnoracial groups. Plasma biomarker cut-off values can vary not only based on analytical variability or ethnoracial variables, but by demographic factors such as age and sex. Thus, these two factors should also be considered when stratifying plasma AD cutoffs.[12,17]

Characterizing plasma biomarkers and their diagnostic precision within Sub-Saharan African (SSA) populations is important, as plasma biomarkers are more cost-effective and easily accessible. Our study aims to provide preliminary RV for plasma protein biomarkers including Aβ42/40 ratio, p-tau181, p-tau217, Nfl, GFAP, IL-1B, IL-10, and TNFα in a sample of adults in the Democratic Republic of Congo with and without dementia to aid in screening, future diagnostic utility, prognostication, and management of AD in the DRC. We hypothesized that in this SSA sample of Congolese, core AD plasma biomarkers concentrations will not be influenced by age or sex. However, we hypothesized that non-specific AD biomarkers show a significant difference across age in this sample. We anticipate that some of these markers, such as p-tau181 or p-tau217, will have potential diagnostic value in this population.

## MATERIALS AND METHODS

### Study population

Participants of this study are community-dwellers from Kinshasa/DRC selected from our prevalence study. [18]Participants were included if they were at least 65 years or older, had a family member or close friend to serve as an informant, and fluent in French or Lingala. We excluded participants who had history of schizophrenia, neurological, or other medical conditions potentially affecting the central nervous system (CNS). To establish neurological status in the absence of established diagnostic criteria for AD in Sub-Saharan Africa (SSA), we screened participants using the Alzheimer’s Questionnaire (AQ)[19] and the Community Screening Instrument for Dementia (CSID)[20]. The AQ was used to assess activities of daily living and symptoms of AD in participants,[19] while the CSID Questionnaire, which is extensively used in many SSA dementia studies,[20] was used to screen cognitive abilities.

Based on cognitive and functional deficits per the Diagnostic and Statistical Manual of Mental Disorders, Fifth Edition, Text Revision (DSM-5-TR) diagnostic criteria,[21] we used CSID cut-offs developed in a study in Brazzaville (Republic of the Congo), the closest city to Kinshasa, to classify participants.[22] Similar to our prior study,[18] participants were classified using CSID and AQ scores (see Figure 1), which yielded 4 groups: major neurocognitive disorder/dementia, mild neurocognitive disorder (MND), subjective cognitive impairment, and healthy control (HC), i.e., normal cognition (Figure 1).

A panel consisting of a neurologist, psychiatrist and neuropsychologist reviewed screening tests, clinical interview, and neurological examination of subjects, of whom 56 were confirmed with a diagnosis of dementia and 58 were considered HC. Of these 114, 29 refused to provide blood samples, leaving 85 participants (75%) in whom plasma biomarkers were obtained (44 dementia and 41 HC). Written informed consent was obtained prior to participants’ undergoing any study procedures. Participants were financially compensated for their time. The procedures were approved by the Ethics Committee/Institutional Review Boards of the University of Kinshasa and Emory University.

**Figure 1.**
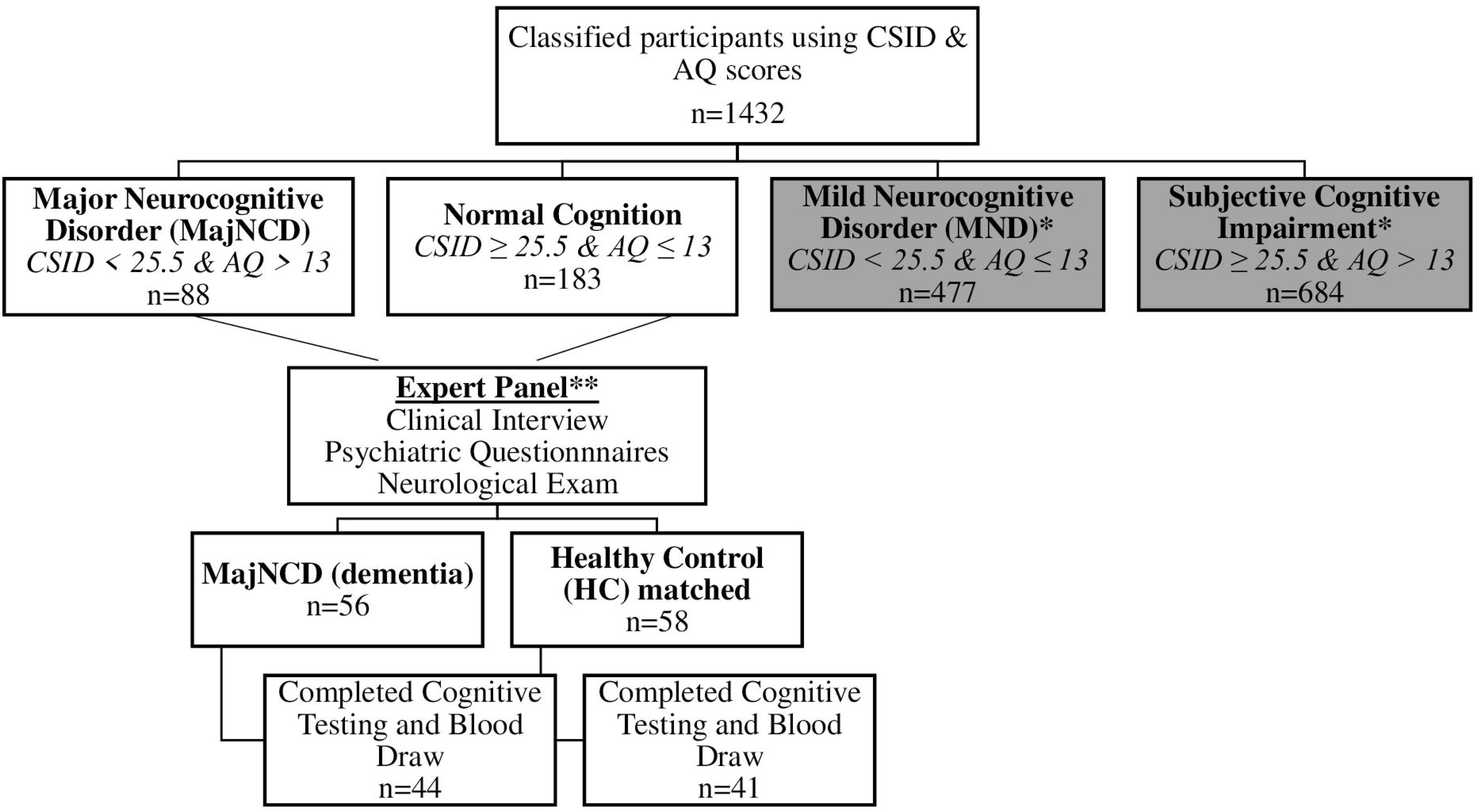
Flow diagram of participant classifications using the CSI-D and the AQ in the current study. **Abbreviations:** CSID (Community Screening Instruments for Dementia); AQ (Alzheimer’s Questionnaire); MajNCD (major neurocognitive disorder); HC (healthy controls); MND (mild neurocognitive disorder).

### Procedure

Qualifying participants answered self-reported questionnaires and underwent cognitive testing alongside standard psychiatric and neurological evaluations to be diagnosed with dementia or to be considered as HC by consensus of an expert panel (neurologist [EE], psychiatrist [GG], and neuropsychologist [JI]). Subjects were interviewed to obtain demographic, socioeconomic, and medical histories and subsequently administered cognitive testing with subtests from the African Neuropsychological Battery (ANB) (cite). Blood samples were obtained at the Medical Center of Kinshasa (CMK) by a phlebotomist. Sample collection protocol and quantification of fluid biomarkers are presented below.

### Measures

### Plasma biomarkers

Blood samples were drawn in the CMK blood laboratory by venipuncture into dipotassium ethylene diamine tetra acetic acid (K_2_ EDTA) tubes. Samples were centrifuged within 15 minutes, and 5 mL of plasma was aliquoted into 0.5 mL polypropylene tubes and stored initially at -20° C for less than a week and then moved to a -80 °C freezer for longer term storage at a CMK laboratory.[16] These aliquots were shipped frozen on dry ice to Emory University. Plasma biomarker concentrations were measured using commercially available Neurology 4-PLEX E (Aβ40, Aβ42, Nfl, and GFAP; lot #503819), P-Tau181 (P-Tau181 v2; lot #503732), IL-1b (lot #503806) and IL-10 (IL-10 2.0, lot #503533) Quanterix kits on the Simoa HD-X platform (Billerica, MA) at UCSF. P-tau217 was measured using the proprietary ALZpath pTau-217 CARe Advantage kit (lot #MAB231122, ALZpath, Inc.) on the Simoa HD-X platform. The instrument operator was blinded to clinical variables. All analytes were measured in duplicate, except for IL-1b, which was measured as a singlicate due to low sample availability. For Aβ40, Aβ42, Nfl, and GFAP, all samples were measured above the lower limit of quantification (LLOQ) of 1.02 pg/mL, 0.378 pg/mL, 0.4 pg/mL and 2.89 pg/mL, respectively. The average coefficient of variation (CV) for Aβ40, Aβ42, Nfl, and GFAP were 6.0%, 6.5%, 5% and 4.6%, respectively. For P-Tau181, all samples were measured above the kit lower limit of quantification (LLOQ) of 0.085 pg/mL, with an average CV of 11.6%. For IL-1b and IL-10, the LLOQ were 0.083 pg/mL and 0.021 pg/mL, respectively. The average CV for IL-10 was 6.1%. For P-tau217 the LLOQ was 0.024 pg/mL and the average CV was 19.8%.

### Statistical Analyses

Statistical analyses were performed using SAS and R statistical software programs. Descriptive statistics for continuous, normally distributed variables are presented as mean ± standard deviation (SD), continuous variables with non-normal distributions are expressed as the median and interquartile range (IQR), and categorical variables are expressed using counts and proportions. Box plots and jittered scatterplots were produced to show the distribution of the plasma biomarkers (the minimum value, the first quartile, the median, the third quartile, and the maximum value), and outliers, overall and also stratified by age and sex. We compared AD biomarkers by age using tertiles for age (50-69 years, 70 -76 years, and 77 years and over). Winsorization of plasma biomarkers to the 95^th^ percentile was used to limit the effect of extreme outliers.

Multiple linear regressions were conducted to evaluate differences in biomarkers by age, sex, and neurological status. Models were adjusted also for education. Logistic regressions were conducted to create receiver operating characteristic (ROC) curve analyses and calculate areas under curve (AUCs) to predict diagnostic accuracy of biomarkers for neurological status (healthy or dementia). Cutoff scores for plasma biomarkers were determined based on optimal sensitivity and specificity for determining the neurological status of having dementia. We used Hosmer and colleagues ROC-AUC categories (Hosmer et al., 2013), which considered the value of <0.600 as “failure,” values between 0.600 and 0.699 as “poor,” values between 0.700 and 0.799 as “fair,” values between 0.800 and 0.899 as “good,” and values 0.900 or greater as “excellent.” We calculated Youden’s indices (sensitivity + specificity – 100) for each plasma biomarker. We selected cutoffs based on the values of the biomarkers that maximized the Youden’s index.

## RESULTS

Demographic data, cognitive scores, clinical data, and plasma biomarker concentrations stratified by neurological status are presented in Table 1. As expected, there were significant differences in cognitive screening scores used in distinguishing neurological status, with healthy individuals having better scores than those with dementia. For clinical data, only HbA1c levels showed a significant difference between HC and dementia, with HC having higher levels of HbA1c than suspected dementia. Diagnostic groups differed in mean levels of Nfl and GFAP after controlling for age and sex (Table 1).

**Table 1.**
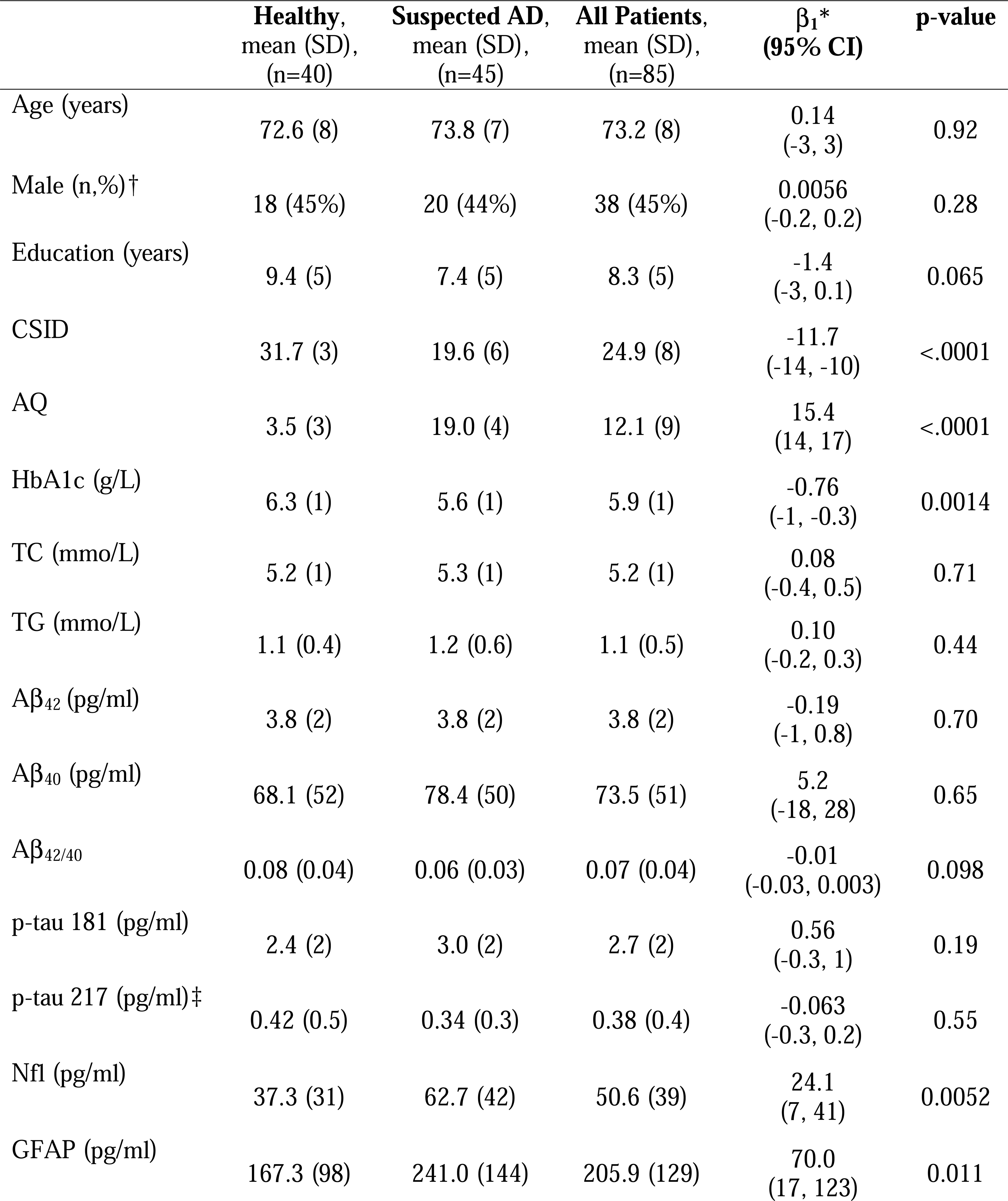
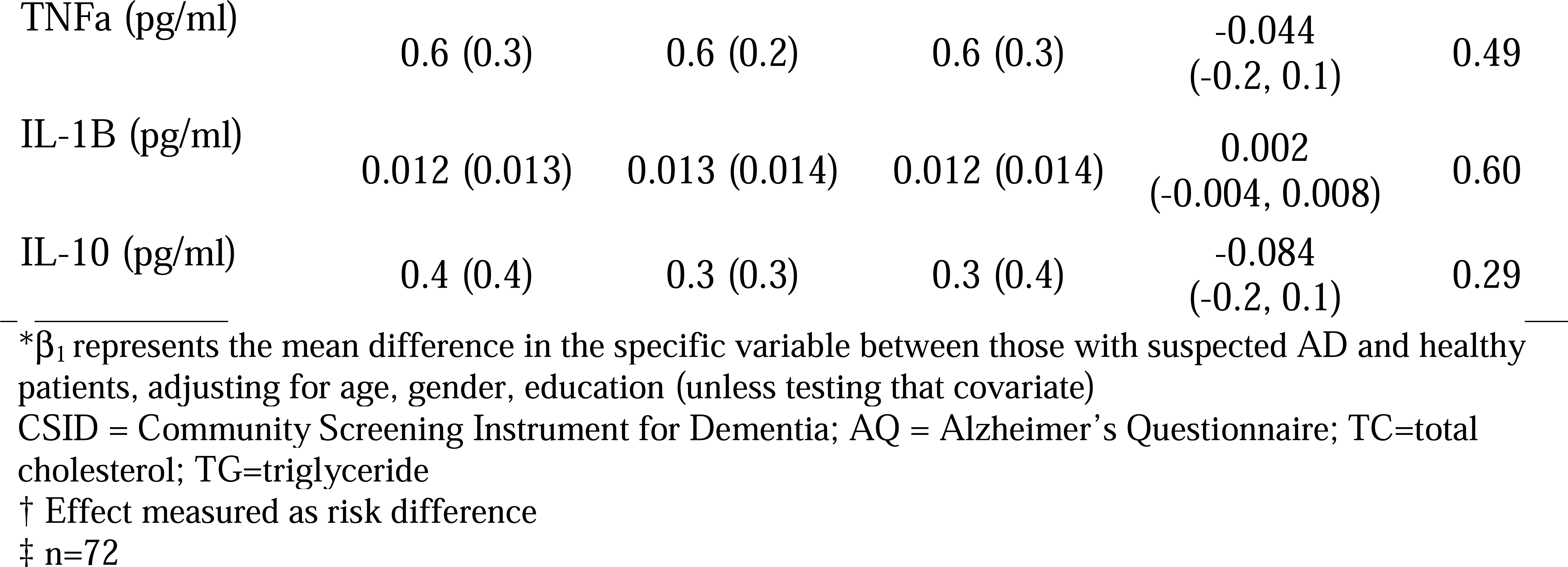
Characteristics of the study sample stratified by cognitive status.

| | Healthy,<br>mean (SD),<br>(n=40) | Suspected AD,<br>mean (SD),<br>(n=45) | All Patients,<br>mean (SD),<br>(n=85) | $\beta_1^*$<br>(95% CI) | p-value |
| --- | --- | --- | --- | --- | --- |
| Age (years) | 72.6 (8) | 73.8 (7) | 73.2 (8) | 0.14<br>(-3, 3) | 0.92 |
| Male (n,%)† | 18 (45%) | 20 (44%) | 38 (45%) | 0.0056<br>(-0.2, 0.2) | 0.28 |
| Education (years) | 9.4 (5) | 7.4 (5) | 8.3 (5) | -1.4<br>(-3, 0.1) | 0.065 |
| CSID | 31.7 (3) | 19.6 (6) | 24.9 (8) | -11.7<br>(-14, -10) | <.0001 |
| AQ | 3.5 (3) | 19.0 (4) | 12.1 (9) | 15.4<br>(14, 17) | <.0001 |
| HbA1c (g/L) | 6.3 (1) | 5.6 (1) | 5.9 (1) | -0.76<br>(-1, -0.3) | 0.0014 |
| TC (mmo/L) | 5.2 (1) | 5.3 (1) | 5.2 (1) | 0.08<br>(-0.4, 0.5) | 0.71 |
| TG (mmo/L) | 1.1 (0.4) | 1.2 (0.6) | 1.1 (0.5) | 0.10<br>(-0.2, 0.3) | 0.44 |
| A$\beta_{42}$ (pg/ml) | 3.8 (2) | 3.8 (2) | 3.8 (2) | -0.19<br>(-1, 0.8) | 0.70 |
| A$\beta_{40}$ (pg/ml) | 68.1 (52) | 78.4 (50) | 73.5 (51) | 5.2<br>(-18, 28) | 0.65 |
| A$\beta_{42/40}$ | 0.08 (0.04) | 0.06 (0.03) | 0.07 (0.04) | -0.01<br>(-0.03, 0.003) | 0.098 |
| p-tau 181 (pg/ml) | 2.4 (2) | 3.0 (2) | 2.7 (2) | 0.56<br>(-0.3, 1) | 0.19 |
| p-tau 217 (pg/ml)‡ | 0.42 (0.5) | 0.34 (0.3) | 0.38 (0.4) | -0.063<br>(-0.3, 0.2) | 0.55 |
| Nfl (pg/ml) | 37.3 (31) | 62.7 (42) | 50.6 (39) | 24.1<br>(7, 41) | 0.0052 |
| GFAP (pg/ml) | 167.3 (98) | 241.0 (144) | 205.9 (129) | 70.0<br>(17, 123) | 0.011 |
| TNFα (pg/ml) | 0.6 (0.3) | 0.6 (0.2) | 0.6 (0.3) | -0.044<br>(-0.2, 0.1) | 0.49 |
| IL-1B (pg/ml) | 0.012 (0.013) | 0.013 (0.014) | 0.012 (0.014) | 0.002<br>(-0.004, 0.008) | 0.60 |
| IL-10 (pg/ml) | 0.4 (0.4) | 0.3 (0.3) | 0.3 (0.4) | -0.084<br>(-0.2, 0.1) | 0.29 |

\* $\beta_1$ represents the mean difference in the specific variable between those with suspected AD and healthy patients, adjusting for age, gender, education (unless testing that covariate)
CSID = Community Screening Instrument for Dementia; AQ = Alzheimer's Questionnaire; TC=total cholesterol; TG=triglyceride
† Effect measured as risk difference
‡ n=72

Figure 2 shows the distribution [minimum, 25^th^ (q1), 50^th^ (q2), 75^th^ (q3), and the maximum], variability, and the skewness of each plasma biomarker. Aβ42, Aβ40, and p-tau 181 are nearly normally distributed, whereas p-tau 217, GFAP, Nfl, TNFα, IL-1b and IL-10 are right skewed.

**Figure 2.**
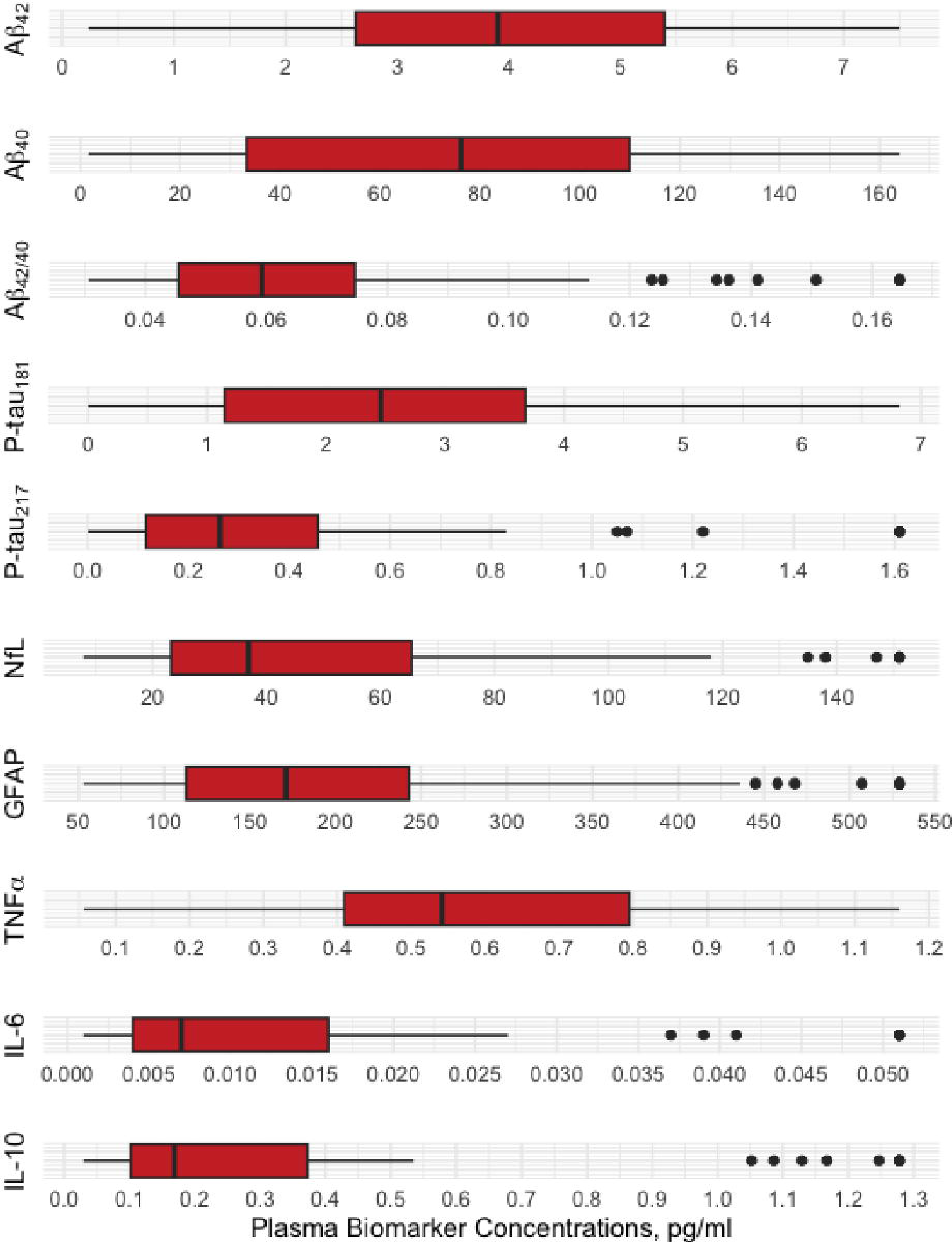
Distribution properties of plasma biomarkers.

The concentrations of plasma biomarkers, except for p-tau181 and GFAP, did not significantly differ by age (Table 2). Plasma p-tau 181 concentrations were significantly higher in participants aged 77 years and older (3.23 (2.12) pg/mL) than in participants aged 50–69 years (1.79 (1.16) pg/mL) or 70–76 years (2.96 (2.32) pg/mL). In addition, plasma GFAP concentrations were significantly higher in participants aged 70-76 years (238.8 (153.7) pg/mL) and 77 years and more (234.0 (115.0) pg/mL) than in participants aged 50–69 years (143.0 (93.5) pg/mL). The concentration of p-tau 217, Nfl, TNFα, and IL-10 were decreased in the age groups of 70-76 years and 77 years and more (Table 2).

**Table 2.** Association of AD biomarkers with age.

| Biomarkers | Age Groups, mean (SD) |  |  | Adjusted Linear Regression |  | Crude Linear Regression |  |
| --- | --- | --- | --- | --- | --- | --- | --- |
| | 50-69 y<br>(n=27) | 70-76 y<br>(n=27) | 77+ y<br>(n=31) | $\beta_1^*$<br>(95% CI) | p-value | $\beta_1$<br>(95% CI) | p-value |
| A$\beta_{42}$ | 3.7 (2) | 3.9 (2) | 3.9 (2) | -0.01<br>(-0.1, 0.1) | 0.78 | 0.002<br>(-0.06, 0.07) | 0.94 |
| A$\beta_{40}$ | 63.3 (39) | 77.2 (53) | 79.0 (58) | 0.41<br>(-1, 2) | 0.61 | 0.88<br>(-0.5, 2) | 0.23 |
| A$\beta_{42/40}$ | 0.07<br>(0.02) | 0.07<br>(0.04) | 0.07<br>(0.04) | 0.0003<br>(-0.00009,<br>0.002) | 0.60 | 0.0001<br>(-0.001,<br>0.001) | 0.85 |
| P-tau<sub>181</sub> | 1.8 (1) | 2.9 (2) | 3.2 (2) | 0.08<br>(0.02, 0.1) | 0.009 | 0.08<br>(0.03, 0.1) | 0.003 |
| P-tau<sub>217</sub> | 0.4 (0.5) | 0.3 (0.4) | 0.4 (0.3) | 0.005<br>(-0.01, 0.02) | 0.49 | 0.0001<br>(-0.01, 0.01) | 0.98 |
| NfI | 48.3 (43) | 46.7 (35) | 56.1 (39) | 0.48<br>(-1, 2) | 0.45 | 0.46<br>(-0.6, 2) | 0.41 |
| GFAP | 143.0<br>(94) | 238.8<br>(154) | 234.0<br>(115) | 4.6<br>(1, 9) | 0.026 | 5.0<br>(2, 9) | 0.005 |
| TNFα | 0.60<br>(0.3) | 0.58<br>(0.3) | 0.62<br>(0.3) | -0.0006<br>(-0.01, 0.009) | 0.90 | -0.0001<br>(-0.008,<br>0.008) | 0.98 |
| IL-1B | 0.01<br>(0.01) | 0.01<br>(0.01) | 0.02<br>(0.02) | 0.0004<br>(-0.00005,<br>0.0008) | 0.084 | 0.0003<br>(-0.0001,<br>0.0006) | 0.17 |
| IL-10 | 0.34<br>(0.4) | 0.30<br>(0.4) | 0.28<br>(0.3) | -0.008<br>(-0.02, 0.003) | 0.15 | -0.004<br>(-0.01, 0.006) | 0.42 |

\* $\beta_1$ represents the mean difference magnitude in biomarker concentrations by 1 year of age increase, adjusting for gender and education

Aβ42/40, p-tau 217 values increase with older age, while Nfl, GFAP and IL-1B values also increase with age. As age group increases, p-tau 181 values increase. There is a rather large spread of values per age group. IL-10 values decrease as age increases (See Figure 3).

**Figure 3.**
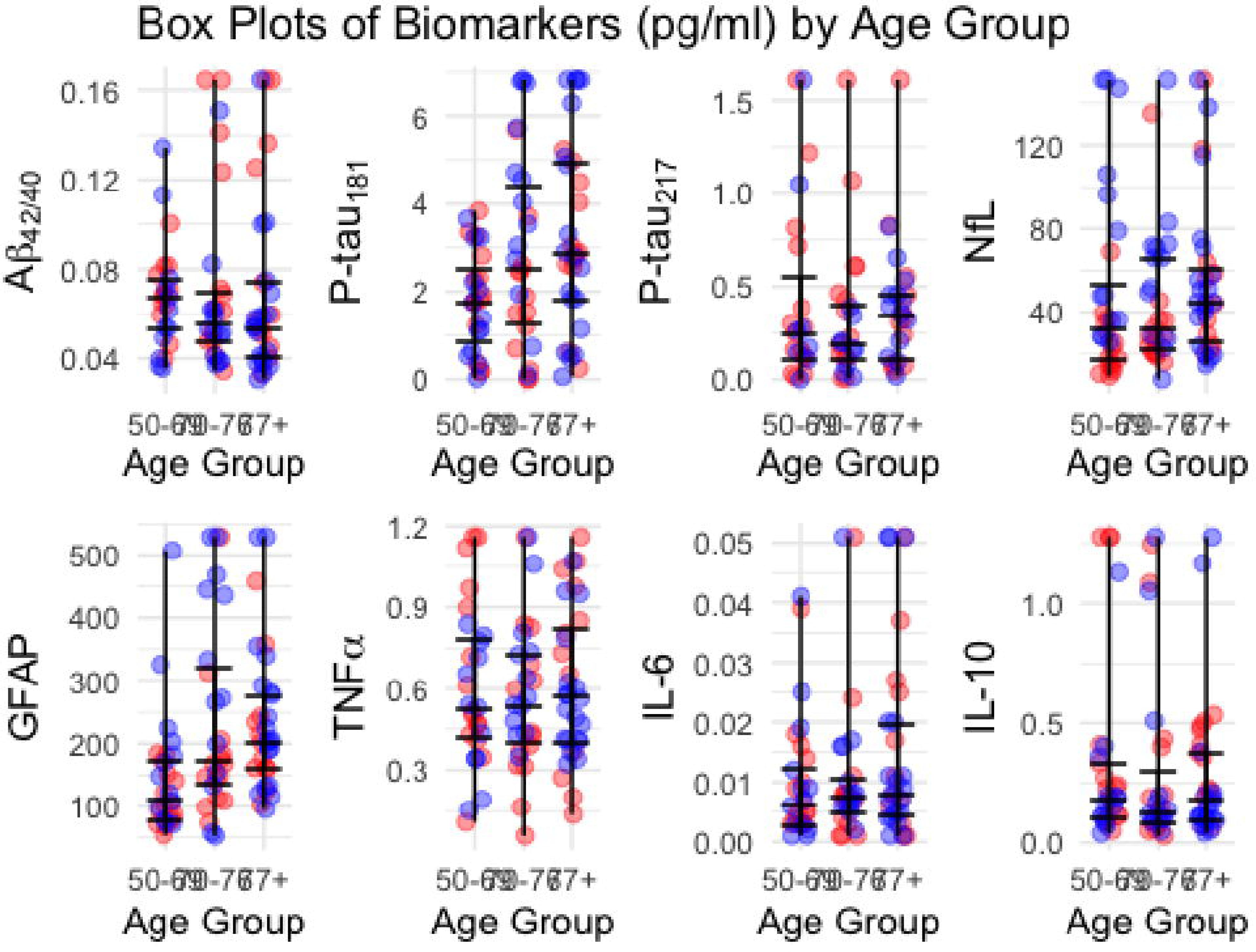
Jitterplots of plasma biomarkers by age group. In these jitter plots, each data point in the form of single dot represents an individual’s biomarker data. The vertical per age group is from q1 (25^th^ quartile) to median to q3 (75^th^ quartile).

Concentrations of plasma biomarkers were not significantly different between men and women (Table 3).

**Table 3.**
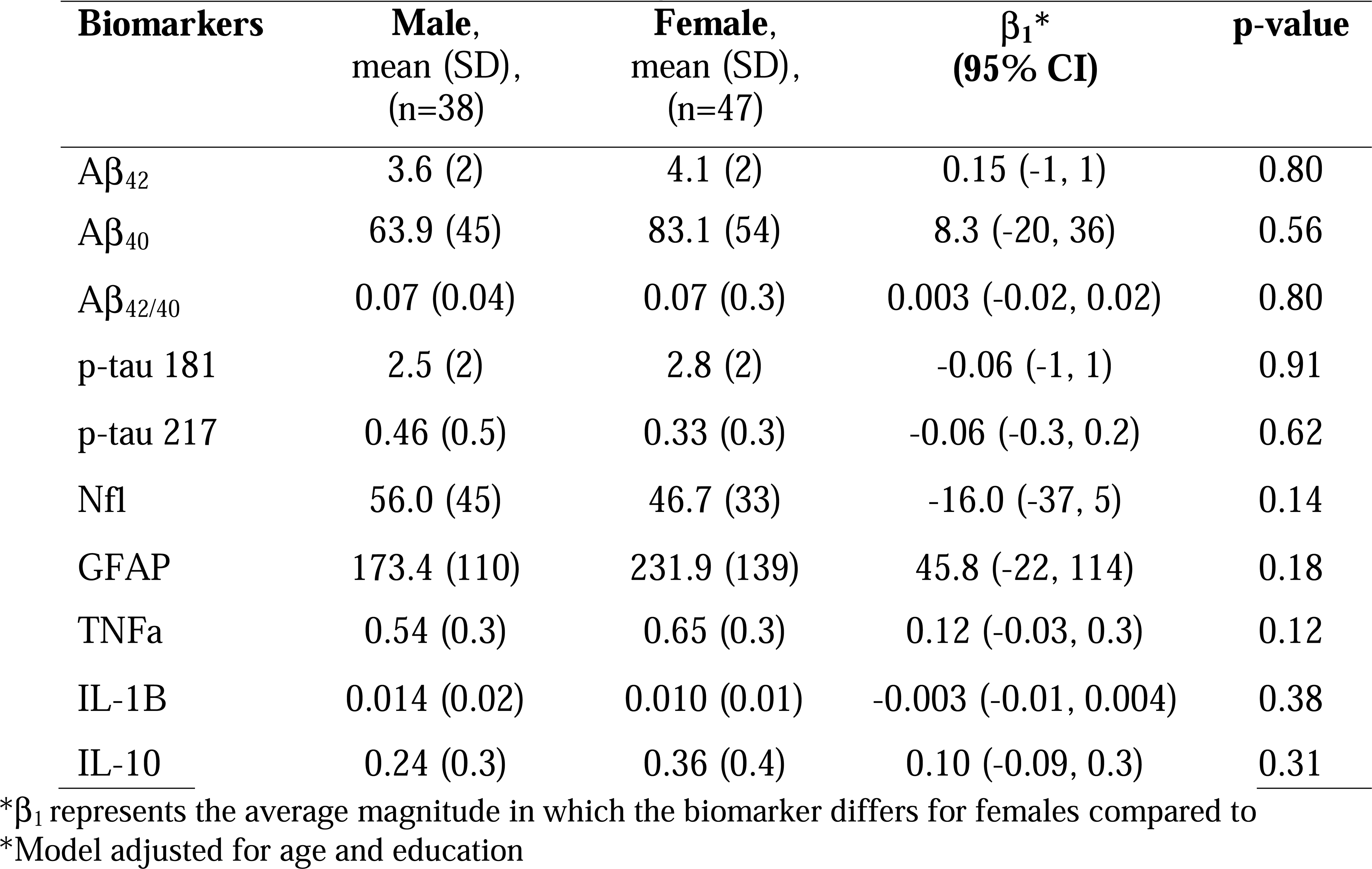
Association of AD biomarkers with gender.

| **Biomarkers** | **Male,**<br>**mean (SD),**<br>**(n=38)** | **Female,**<br>**mean (SD),**<br>**(n=47)** | **$\beta_1^*$**<br>**(95% CI)** | **p-value** |
| --- | --- | --- | --- | --- |
| A$\beta_{42}$ | 3.6 (2) | 4.1 (2) | 0.15 (-1, 1) | 0.80 |
| A$\beta_{40}$ | 63.9 (45) | 83.1 (54) | 8.3 (-20, 36) | 0.56 |
| A$\beta_{42/40}$ | 0.07 (0.04) | 0.07 (0.3) | 0.003 (-0.02, 0.02) | 0.80 |
| p-tau 181 | 2.5 (2) | 2.8 (2) | -0.06 (-1, 1) | 0.91 |
| p-tau 217 | 0.46 (0.5) | 0.33 (0.3) | -0.06 (-0.3, 0.2) | 0.62 |
| NfI | 56.0 (45) | 46.7 (33) | -16.0 (-37, 5) | 0.14 |
| GFAP | 173.4 (110) | 231.9 (139) | 45.8 (-22, 114) | 0.18 |
| TNFα | 0.54 (0.3) | 0.65 (0.3) | 0.12 (-0.03, 0.3) | 0.12 |
| IL-1B | 0.014 (0.02) | 0.010 (0.01) | -0.003 (-0.01, 0.004) | 0.38 |
| IL-10 | 0.24 (0.3) | 0.36 (0.4) | 0.10 (-0.09, 0.3) | 0.31 |

\* $\beta_1$ represents the average magnitude in which the biomarker differs for females compared to
\*Model adjusted for age and education

The distribution of biomarker concentrations by sex is presented in Figure 4. Biomarker concentrations followed a normal distribution or were positively skewed.

**Figure 4.**
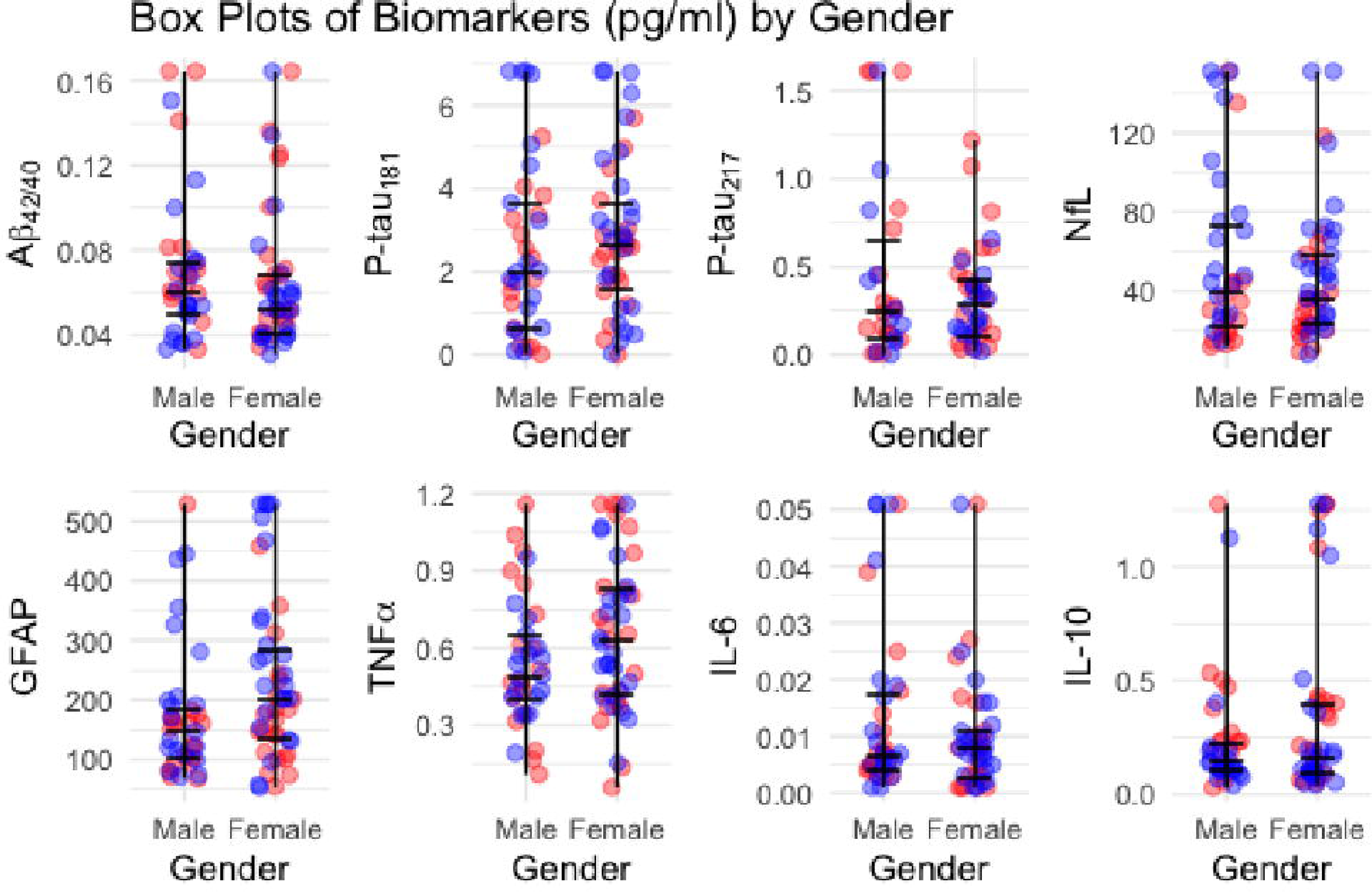
Jitter plots of plasma biomarkers by sex.

Table 4 displays the AUC values for prediction of dementia based on plasma biomarkers. AUC ranges fell between 0.64-0.74 (95% *CIs* ranging from 0.52 – 0.86). TNFα, IL-1b and IL-10 had higher sensitivity than other plasma biomarkers, followed by GFAP. P-tau 217 [0.74 (0.61, 0.86)], GFAP [0.72 (0.61, 0.83), and Nfl [0.71 (0.60, 0.82)] had the highest AUC values.

**Table 4.** Sensitivity, specificity, and overall discrimination (AUC) of plasma biomarkers in distinguishing dementia status.

| **Biomarker** | **Cutoff** | **Sensitivity /**<br>**Specificity** | **AUC (95% CI)** |
| --- | --- | --- | --- |
| Aβ<sub>42/40</sub> | 0.061 | 73.0 / 47.7 | 0.68 (0.56, 0.80) |
| p-Tau 181 | 4.50 | 63.2 / 53.3 | 0.67 (0.55, 0.78) |
| p-Tau 217 | 0.008 | 70.0 / 66.7 | 0.72 (0.59, 0.84) |
| Nfl | 36.5 | 52.5 / 80.0 | 0.73 (0.62, 0.84) |
| GFAP | 176 | 67.5 / 64.4 | 0.72 (0.61, 0.83) |
| TNFα | 1.16 | 67.5 / 47.7 | 0.64 (0.52, 0.76) |
| IL_1b | 0.011 | 69.2 / 55.6 | 0.65 (0.52, 0.77) |
| IL_10 | 0.38 | 80.0 / 40.0 | 0.65 (0.54, 0.77) |

## DISCUSSION

The revised and updated AA criteria have brought major changes to the diagnosis of AD dementia from a purely cognitive diagnosis to a biological clinical diagnostic algorithmic approach.[3] The presence of CSF or plasma amyloid and p-tau 181 or 217 (mostly p-tau 217) has been considered as sensitive and specific to AD with Nfl and GFAP as important non-specific AD biomarkers. The current study has provided preliminary RVs for plasma protein biomarkers, including Aβ42/40 ratio, p-tau181, p-tau217, Nfl, and GFAP, IL-1B, IL-10, and TNFα in a sample of adults in the DRC with and without dementia.

This research explores the clinical performance of established plasma AD and neurodegeneration biomarkers in an African population for which there are no previous biomarker data. Our first hypothesis was partially supported, as age groups did not significantly differ in core AD biomarkers and non-inflammatory/immune AD biomarkers except for p-tau 181 across ages in this SSA sample of Congolese adults. In contrast, non-specific AD biomarkers showed significant age differences, aside from GFAP. This is similar to a recent study comparing biomarker levels (plasma Aβ42/40, p-tau181, GFAP, and Nfl) in participants who were cognitively unimpaired, mildly cognitively impaired, and Aβ-PET–positive participants across the AD pathology continuum.[15] Whereas, in the AD group, Aβ42/40, p-tau181, and GFAP did not show a significant difference across ages, Nfl was shown to correlate with age.[15]

As we predicted, there were no significant differences between women and men in all plasma biomarkers in this sample. Similar findings were reported by a Canadian population-based cohort which also found no significant sex differences in plasma AD biomarkers.[17] Similarly, in a sample of healthy Chinese, Chen and colleagues did not find significant sex differences in plasma Aβ42, Aβ40, or Aβ42/40 ratio; however, men had greater plasma p-tau181, p-tau181/t-tau ratio, and p-tau181/Aβ42 ratio than women.[16]

One interesting finding is the lack of predictive abilities of core plasma biomarkers to classify subjects as having AD pathology. We found a difference between clinical diagnosis based on cognitive tests and diagnostic classification based on AD core plasma biomarkers (Aβ42/40 and p-tau). However, non-specific biomarkers (e.g., Nfl and GFAP) and p-tau 217 had good AUC. Pais and colleagues also found discrepancies between cognitive decline and the diagnostic classification based on AD biomarkers in many studies.[14] Future research should continue to explore the predictive value of plasma biomarkers in various ethnically and culturally diverse samples. Prior research has shown variation in cutoffs by ethnic group. A review by Pais and colleagues (2023) found that plasma AD biomarker cut-off values can vary not only based on analytical variability, but by demographic factors, which can explain the variation of plasma biomarkers across ethnoracial groups, highlighting the importance of studying the underlying pathophysiology in these groups.[12,14,17] Thus, in-house cut-offs may better represent specific populations with the understanding that they are assay-dependent.[14,15] Establishment of clear cut-off criteria is important for potential future clinical utility.

This study is the first in the SSA to attempt to provide preliminary RVs for core and non-specific AD plasma protein biomarkers in a sample of adults in the DRC with and without dementia. Our findings should be interpreted considering several limitations, such as the cross-sectional nature of the study, limited sample size, and lack of amyloid PET imaging or CSF biomarker measurements confirming AD pathology. These RVs should be further validated in longitudinal studies with larger sample size. Furthermore, this is the first study in the Congo where the population can be phenotyped in biofluids. Future research could benefit from sending the same samples for plasma p-tau217 via LabCorp or C2N (which are commercially available and have established cut points), to compare the performance. The screening measures used (CSID and AQ) have not been validated in the SSA/DRC. To that extent, there have been recent studies looking at these relationships with more commonly used cognitive screeners, such as the Mini Mental Status Exam (MMSE) and Montreal Cognitive Assessment (MoCA) across the globe and in different diagnostic groups.[23,24] This study included only subjects with suspected dementia and healthy controls. Those with cognitive difficulties seen in between these two categories (e.g., MCI, subjective memory complaints) were excluded, leaving only the extremes of the dementia spectrum. Future studies should conduct statistical analyses across all 4 groups (healthy controls, MCI, subjective memory complaint and dementia). In the same vein, this study did not characterize the etiology of the dementia syndrome. AD biomarker performance is best in amnestic or logopenic phenotypes. Thus, if our sample had a mixture of executive, behavioral, or mixed phenotypes, the diagnostic accuracy of the plasma AD biomarkers would be compromised. For example, plasma AD biomarkers correlate well with measures of verbal or visuospatial memory or screening tests that rely heavily on memory (e.g., MMSE, MoCA).[23–26] Future studies should also aim to replicate our findings using AD biomarkers in CSF, amyloid or tau brain PET, or mass spectrometry of plasma biomarkers. Additionally, Simoa has limitations for the measurements of plasma AD biomarkers.[14] Thus, continued investigation into racial differences in AD biomarkers and relation to AD-dementia using these gold standard techniques (e.g., brain amyloid PET, CSF) should be conducted. Finally, the findings of this study are exploratory, and we caution that evaluation of these biomarkers in novel populations to support clinical assessments may not be as straightforward as expected. Replication of findings on large scale sample of controls is warranted.

## AUTHOR CONTRIBUTIONS

JI: Visualization, Validation, Supervision, Software, Resources, Project administration, Methodology, Investigation, Funding acquisition, Formal analysis, Data curation, Conceptualization, Writing – review & editing, Writing – original draft. KJ: Writing – review & editing. PM: Writing – review & editing. SSP: Writing – review & editing, Writing – original draft. MS: Writing – review & editing, Writing – original draft. EE: Writing – review & editing. GG: Writing – review & editing. NT: Writing – review & editing. IK: Writing – review & editing. SM: Writing – review & editing. LM: Writing – review & editing. CT: Writing – review & editing. AS: Writing – review & editing. JR: Writing – review & editing. BC: Writing – review & editing. AL: Writing – review & editing. JK: Writing – review & editing. AB: Writing – review & editing. AJ: Writing – review & editing. AG: Writing – review & editing. AA: Writing – review & editing.

## Data Availability

All data produced in the present study are available upon reasonable request to the authors

## ACKNOWLEDGEMENTS

The authors have no acknowledgments to report.

## REFERENCES

[1] Dubois B, Villain N, Frisoni GB, et al. Clinical diagnosis of Alzheimer’s disease: recommendations of the International Working Group. Lancet Neurol. 2021;20(6):484–496. doi:10.1016/S1474-4422(21)00066-1

[2] Blennow K, Zetterberg H. Biomarkers for Alzheimer’s disease: current status and prospects for the future. J Intern Med. 2018;284(6):643–663. doi:10.1111/joim.12816

[3] Babić Leko M, Nikolac Perković M, Klepac N, et al. IL-1β, IL-1B, IL-10, and TNFα Single Nucleotide Polymorphisms in Human INfluence the Susceptibility to Alzheimer’s Disease Pathology. J Alzheimers Dis JAD. 2020;75(3):1029-1047. doi:10.3233/JAD-200056

[4] Giacomucci G, Mazzeo S, Bagnoli S, et al. Plasma neurofilament light chain as a biomarker of Alzheimer’s disease in Subjective Cognitive Decline and Mild Cognitive Impairment. J Neurol. 2022;269(8):4270–4280. doi:10.1007/s00415-022-11055-5

[5] Gulisano W, Maugeri D, Baltrons MA, et al. Role of Amyloid-β and Tau Proteins in Alzheimer’s Disease: Confuting the Amyloid Cascade. J Alzheimers Dis JAD. 2018;64(s1):611–631. doi:10.3233/JAD-179935

[6] Kim KY, Shin KY, Chang KA. GFAP as a Potential Biomarker for Alzheimer’s Disease: A Systematic Review and Meta-Analysis. Cells. 2023;12(9):1309. doi:10.3390/cells12091309

[7] Silva NML e, Goncalves RA, Pascoal TA, et al. Pro-iNflammatory interleukin-6 signaling links cognitive impairments and peripheral metabolic alterations in Alzheimer’s disease. Transl Psychiatry. 2021;11(1):251–251. doi:10.1038/s41398-021-01349-z

[8] Pais MV, Forlenza OV, Diniz BS. Plasma Biomarkers of Alzheimer’s Disease: A Review of Available Assays, Recent Developments, and Implications for Clinical Practice. J Alzheimers Dis Rep. 2023;7(1):355–380. doi:10.3233/ADR-230029

[9] Bishop GM, Robinson SR. Physiological roles of amyloid-beta and implications for its removal in Alzheimer’s disease. Drugs Aging. 2004;21(10):621–630. doi:10.2165/00002512-200421100-00001

[10] Small DH, McLean CA. Alzheimer’s disease and the amyloid β protein : What is the role of amyloid? J Neurochem. 1999;73(2):443–449. doi:10.1046/j.1471-4159.1999.0730443.x

[11] Constantinides VC, Paraskevas GP, Boufidou F, et al. CSF Aβ42 and Aβ42/Aβ40 Ratio in Alzheimer’s Disease and Frontotemporal Dementias. 2023;13(4):783. doi:10.3390/diagnostics13040783

[12] Lewczuk P, Matzen A, Blennow K, et al. Cerebrospinal Fluid Aβ42/40 Corresponds Better than Aβ42 to Amyloid PET in Alzheimer’s Disease. J Alzheimer’s Dis. 2017;55(2):813–822. doi:10.3233/JAD-160722

[13] Janelidze S, Teunissen CE, Zetterberg H, et al. Head-to-Head Comparison of 8 Plasma Amyloid-β 42/40 Assays in Alzheimer Disease. Arch Neurol Chic. 2021;78(11):1375–1382. doi:10.1001/jamaneurol.2021.3180

[14] Avila J, Lucas JJ, Perez M, Hernandez F. Role of tau protein in both physiological and pathological conditions. Physiol Rev. 2004;84(2):361–384. doi:10.1152/physrev.00024.2003

[15] Brion JP, Anderton BH, Authelet M, et al. Neurofibrillary tangles and tau phosphorylation. Biochem Soc Symp. 2001;67:81-88. doi:10.1042/bss0670081

[16] Wattmo C, Blennow K, Hansson O. Cerebro-spinal fluid biomarker levels: phosphorylated tau (T) and total tau (N) as markers for rate of progression in Alzheimer’s disease. BMC Neurol. 2020;20(1):10–10. doi:10.1186/s12883-019-1591-0

[17] Thijssen EH, Joie R, Strom A, et al Advancing Research and Treatment for Frontotemporal Lobar Degeneration investigators (2021). Plasma phosphorylated tau 217 and phosphorylated tau 181 as biomarkers in Alzheimer’s disease and frontotemporal lobar degeneration: a retrospective diagnostic performance study. Lancet Neurol. 20(9):739-752. doi:10.1016/S1474-4422(21)00214-3

[18] Mielke MM, Syrjanen JA, Blennow K, et al. Plasma and CSF neurofilament light: Relation to longitudinal neuroimaging and cognitive measures. Neurology. 2019;93(3):252–260. doi:10.1212/WNL.0000000000007767

[19] Wei DC, Morrison EH. Histology, Astrocytes. In: StatPearls. StatPearls Publishing; 2023.

[20] Hansson O, Edelmayer RM, Boxer AL, et al. The Alzheimer’s Association appropriate use recommendations for blood biomarkers in Alzheimer’s disease. Alzheimers Dement J Alzheimers Assoc. 2022;18(12):2669–2686. doi:10.1002/alz.12756

[21] Pereira JB, Janelidze S, Smith R, et al. Plasma GFAP is an early marker of amyloid-β but not tau pathology in Alzheimer’s disease. Brain J Neurol. 2021;144(11):3505–3516. doi:10.1093/brain/awab223

[22] Arosio B, Trabattoni D, Galimberti L, et al. Interleukin-10 and interleukin-6 gene polymorphisms as risk factors for Alzheimer’s disease. Neurobiol Aging. 2004;25(8):1009–1015. doi:10.1016/j.neurobiolaging.2003.10.009

[23] Windon C, Iaccarino L, Mundada N, et al. Comparison of plasma and CSF biomarkers across ethnoracial groups in the ADNI. Alzheimers Dement Diagn Assess Dis Monit. 2022;14(1):e12315. doi:10.1002/dad2.12315

[24] Meeker KL, Ances B, Petersen M, et al. Effects of structural and social determinants of health and comorbidities on ethno-racial differences in ATN plasma biomarkers of Alzheimer’s disease: A HABS-HD study. Alzheimers Dement. 2023;19(S14):e078605. doi:10.1002/alz.078605

[25] Hall JR, Petersen M, Johnson L, O’Bryant SE. Characterizing Plasma Biomarkers of Alzheimer’s in a Diverse Community-Based Cohort: A Cross-Sectional Study of the HAB-HD Cohort. Front Neurol. 2022;13:871947. doi:10.3389/fneur.2022.871947

[26] Hajjar I, Yang Z, Okafor M, et al. Association of Plasma and Cerebrospinal Fluid Alzheimer Disease Biomarkers With Race and the Role of Genetic Ancestry, Vascular Comorbidities, and Neighborhood Factors. JAMA Netw Open. 2022;5(10):e2235068. doi:10.1001/jamanetworkopen.2022.35068

[27] Deniz K, Ho CCG, Malphrus KG, et al. Plasma Biomarkers of Alzheimer’s Disease in African Americans. J Alzheimers Dis. 2021;79(1):323–334. doi:10.3233/JAD-200828

[28] Schindler SE, Karikari TK, Ashton NJ, et al. Effect of Race on Prediction of Brain Amyloidosis by Plasma Aβ42/Aβ40, Phosphorylated Tau, and Neurofilament Light. Neurology. 2022;99(3):e245–e257. doi:10.1212/WNL.0000000000200358

[29] Boots EA, Feinstein DL, Leurgans S, et al. Acute versus chronic iNflammatory markers and cognition in older black adults: Results from the Minority Aging Research Study. Brain Behav Immun. 2022;103:163–170. doi:10.1016/j.bbi.2022.04.014

[30] Wang T, Xiao S, Liu Y, et al. The efficacy of plasma biomarkers in early diagnosis of Alzheimer’s disease. Int J Geriatr Psychiatry. 2014;29(7):713–719. doi:10.1002/gps.4053

[31] Altomare D, Stampacchia S, Ribaldi F, et al. Plasma biomarkers for Alzheimer’s disease: a field-test in a memory clinic. J Neurol Neurosurg Psychiatry. 2023;94(6):420–427. doi:10.1136/jnnp-2022-330619

[32] Chatterjee P, Pedrini S, Ashton NJ, et al. Diagnostic and prognostic plasma biomarkers for preclinical Alzheimer’s disease. Alzheimers Dement. 2022;18(6):1141–1154. doi:10.1002/alz.12447

[33] Cooper JG, Stukas S, Ghodsi M, et al. Age specific reference intervals for plasma biomarkers of neurodegeneration and neurotrauma in a Canadian population. Clin Biochem. 2023;121–122:110680. doi:10.1016/j.clinbiochem.2023.110680

[34] Mielke MM. Consideration of Sex Differences in the Measurement and Interpretation of Alzheimer’s Disease-Related Biofluid-Based Biomarkers. J Appl Lab Med. 2020;5(1):158–169. doi:10.1373/jalm.2019.030023

[35] Honig LS, Kang MS, Lee AJ, et al. Evaluation of Plasma Biomarkers for A/T/N Classification of Alzheimer Disease Among Adults of Caribbean Hispanic Ethnicity. JAMA Netw Open. 2023;6(4). doi:10.1001/jamanetworkopen.2023.8214

[36] Chen J, Zhao X, Zhang W, et al. Reference intervals for plasma amyloid-β, total tau, and phosphorylated tau181 in healthy elderly Chinese individuals without cognitive impairment. Alzheimers Res Ther. 2023;15(1):100. doi:10.1186/s13195-023-01246-1

[37] Prince M, Guerchet M, Prina M. The Global Impact of Dementia 2013-2050. Alzheimer’s Disease International; 2013.

[38] Ikanga J, Patel SS, Roberts BR, et al. Association of plasma biomarkers with cognitive function in persons with dementia and cognitively healthy in the Democratic Republic of Congo. Alzheimers Dement Diagn Assess Dis Monit. 2023;15(4):e12496. doi:10.1002/dad2.12496

[39] Schwinne M, Alonso A, Roberts BR, et al. The Association of Alzheimer’s Disease-related Blood-based Biomarkers with Cognitive Screening Test Performance in the Congolese Population in Kinshasa. medRxiv. Published online October 31, 2023:2023.08.28.23294740. doi:10.1101/2023.08.28.23294740

[40] Bucci M, Bluma M, Savitcheva I, et al. Profiling of plasma biomarkers in the context of memory assessment in a tertiary memory clinic. Transl Psychiatry. 2023;13. doi:10.1038/s41398-023-02558-4

[41] Ashton NJ, Brum WS, Molfetta G, et al. Diagnostic accuracy of the plasma ALZpath pTau217 immunoassay to identify Alzheimer’s disease pathology. Published online 2023. doi:10.1101/2023.07.11.23292493

[41] Quest Diagnostics: Test Directory. Published online October 2023. https://testdirectory.questdiagnostics.com/test/test-guides/TS_AD_Detect_BetaRatioPlasma/quest-ad-detect

